# The influence of climate factors on COVID-19 transmission in Malaysia: An autoregressive integrated moving average (ARIMA) model

**DOI:** 10.1101/2020.08.14.20175372

**Authors:** Cho Naing, Han Ni, Htar Htar Aung, Elaine Chan, Joon Wah Mak

## Abstract

**Background:** A unique concern pertaining to the spread of COVID-19 across countries is the asymmetry of risk and the irrational fear of a new pandemic and its possible serious consequences. This study aimed to perform a time series analysis on the association between climate factors and daily cases of COVID-19 in Malaysia up to 15 July 2020. The second objective was to predict daily new cases using a forecasting technique. To address within-country variations, the analysis was extended to the state level with Sarawak state as an example.

**Methodology/Principal Findings:** Datasets on the daily confirmed cases and climate variables in Malaysia and Sarawak state were obtained from publicly accessible official websites. A descriptive analysis was performed to characterize all the important variables over the study period. An autoregressive integrated moving average (ARIMA) model was introduced using daily cases as the dependent variable and climate parameters as the explanatory variables.

For Malaysia, the findings suggest that, *ceteris paribus*, the number of COVID-19 cases decreased with increasing average temperature (p=0.003) or wind speed (p=0.029). However, none of the climate parameters showed a significant relationship with the number of COVID-19 cases in Sarawak state. Forecasts from the ARIMA models showed that new daily COVID-19 cases had already reached the outbreak level and a decreasing trend in both settings. Holding other parameters constant, a small number of new cases (approximately a single digit) is a probable second wave in Sarawak state,

**Conclusions/Significance:** The findings suggest that climate parameters and forecasts are helpful for reducing the uncertainty in the severity of future COVID-19 transmission. A highlight is that forecasts will be a useful tool for making decisions and taking the appropriate interventions to contain the spread of the virus in the community.

## Introduction

As of 15 June 2020, 7,823,291 confirmed cases and 431,541 deaths from the novel coronavirus disease (COVID-19) have been reported from the WHO’s six regions [1]. In Malaysia, the first sporadic case was detected on 24thJanuary 2020, and as of 16 July 2020, 8,739 confirmed cases and 122 deaths have been reported [2]. A unique concern across countries is the asymmetry of risk and the irrational fear of a new pandemic with its possible catastrophic consequences, as documented with the 1918 Spanish flu that killed an estimated 50 million people worldwide. To reduce uncertainty, governments need to accurately forecast the spread of confirmed cases. According to current evidence, the COVID-19 virus ((severe acute respiratory syndrome coronavirus 2 (SARS-CoV-2)) is primarily transmitted among people through respiratory droplets and contact routes [3]. As such, climate factors such as temperature and/or relative humidity may influence the transmission of coronavirus [4]- by affecting its survival. Previously, studies have documented the relationship between weather parameters and severe acute respiratory coronavirus (SARS-CoV) and Middle East respiratory syndrome coronavirus (MERS-CoV).

Time series studies are most relevant when data have been accumulated over a considerably long period of time. A common challenge to time series methodology is the availability of data. We were able to identify the weather parameters and daily COVID-19 cases at the country level instead of the weekly, monthly or annual aggregates. This finer temporal resolution will allow us to further characterize short-period variations. Despite the sustained transmission of COVID-19 cases, the effects of climate factors on the population risk of COVID-19 in Malaysia have not yet been assessed in detail.

Hence, two research questions arise: *“Is there a role of climate parameters on the possible viral spread in Malaysia?”* and *“what is a forecast of regional transmission of new cases?”*. Taken together, the objective of the present study was to perform a time series analysis on the association between climate factors and daily cases of COVID-19 in Malaysia between January 2020 and 15 July 2020. The second objective was to predict daily new cases using a forecasting technique. Further, to address within-country variations, we extend our analysis to the state level with Sarawak state as an example.

We anticipate that our findings will be useful not only for policymakers in Malaysia but also for countries that will experience inclement weather conditions in 2020 to prepare the necessary public health response for the potential spread of COVID-19.

## Materials and Methods

We performed the present study following a prior protocol. A description of the protocol is available from the corresponding author on request.

### Study area

Malaysia is located between 2° and 7° N, bordering Thailand to the north and Singapore to the south. Sarawak state (1.5533°N, 110.3592°E) is an administrative region of Malaysia located on Borneo Island.

### Data collection

#### COVID-19 incidence

Publicly accessible datasets of the daily confirmed cases and deaths in Malaysia were obtained from the official website of the Malaysian Ministry of Health (MOH). These data were cross-checked with the John Hopkins University Coronavirus Resource Center Repository (https://data.humdata.org/dataset/novel-coronavirus-2019-ncov-cases) up to 15th July 2020

These data originated from the active and passive case surveillance system in Malaysia. We extracted the total daily counts of confirmed cases up to 15th July 2020. The first COVID-19 case was announced in China in late December 2019, and the first case in Malaysia was announced on 23^rd^ January 2020. This finding implies that COVID-19 had spread to Malaysia within one month.

#### Climate factors

The daily mean temperature (°F), relative humidity (%), precipitation (%) and wind velocity (mph) over the study period were collected from an archive of weather data for Malaysia (https://weatherspark.com/history/). For the country level, we used the means of the climate parameters across the reported states.

### Statistical analysis

First, a descriptive analysis was performed to characterize all the variables over the study period in Malaysia and Sarawak state. Then, we introduced an autoregressive integrated moving average (ARIMA) model, using daily cases as the dependent variable and the climate parameters as the explanatory variables.

A structural ARIMA model was introduced to examine the association between the number of COVID-19 cases (daily counts) and the various explanatory variables (i.e., the mean temperature, wind speed, precipitation and relative humidity). We computed various permutations of the order of the autoregressive (AR), order of integration (I) and order of moving average (MA) terms and chose the optimal combination of parameters using the mean squared error. The correlograms were visualized to help in determining the orders of the MA and AR terms to include in the model. Before modeling the daily confirmed cases, we checked the stationarity of the data series by the augmented Dickey-Fuller (ADF) test [5]. A model was first applied to quantify the aggregate country-level associations between the climate variables and the daily cases of COVID-19. Another model was applied to quantify the state-specific association for Sarawak state, Malaysia.

We applied the *auto.arima(1,1,0)* function to forecast new cases. The best order of an ARIMA process was determined through a unit root test to identify the appropriate degree of differencing and by the minimization of the corrected Akaike information criterion (AIC) [5]. We then conducted fourteen-day-ahead point forecasts and calculated the prediction intervals, taking into account the incubation period. Two analytical models were introduced depending on the situation of COVID-19 in Malaysia in general and Sarawak state in particular.

The forecast accuracy was checked with four accuracy measures: the mean absolute error (MAE), mean absolute percentage error (MAPE), mean absolute scaled error (MASE), and root mean square error (RMSE) [6]. We performed the Ljung–Box test, for which a p-value greater than 0.05 means that the residuals for the current time series model are independent [5].

The basic ARIMA forecasting estimated equation is

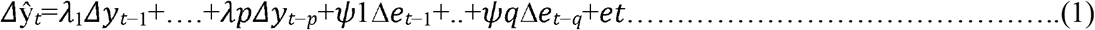

where

= degree of nonseasonal differences,
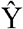= time series that will be predicted at time t,
P = lag order of AR,
Λ= the coefficient of each parameter,
Q = lag order of MA,
Ψ= coefficient of each parameter q, and
*et*= the residuals of the errors at time t.

STATA 16.1 (StataCorp, Texas, USA) was used to calculate the descriptive statistics, and the “*Forecast*” package in *R* (R Core team) was used to forecast the confirmed COVID-19 cases.

## Results

Table 1 presents the characteristics of the variables for Malaysia and Sarawak state.

**Table 1.**
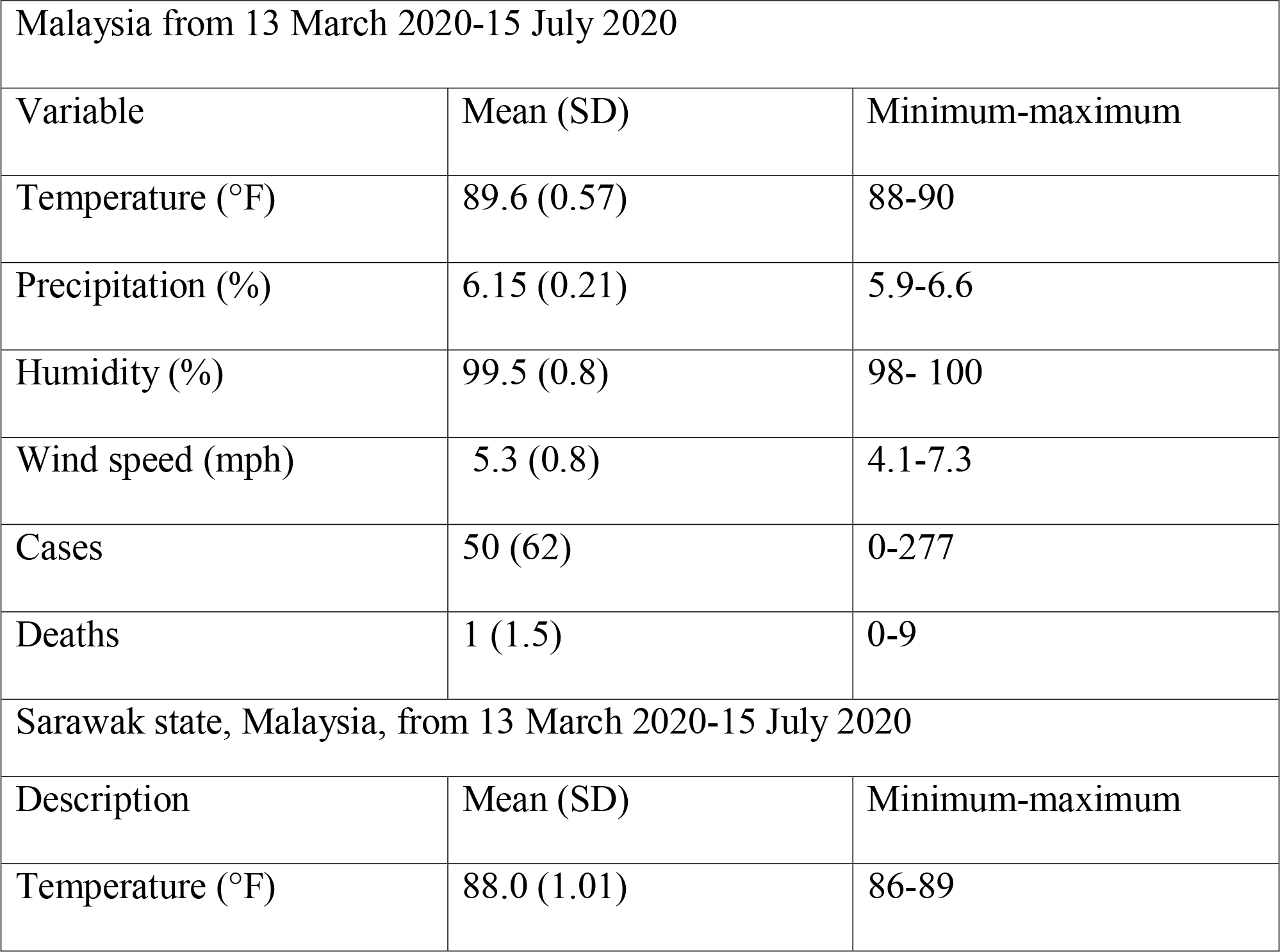
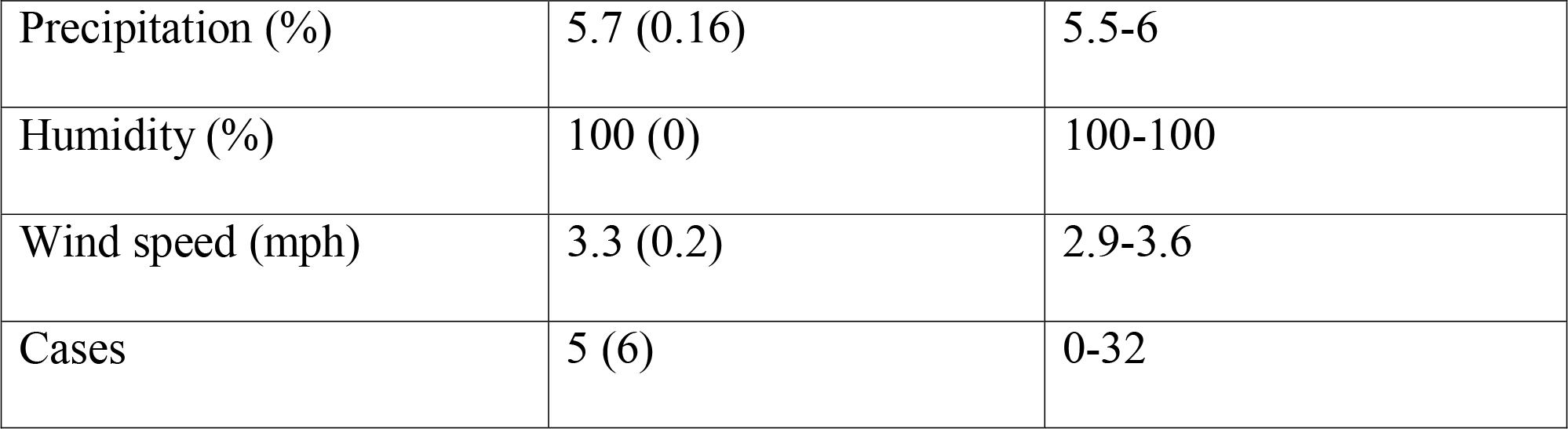
Characteristics of the weather variables and new reported cases

During the period from 24^th^ January 2020 to 15^th^ July 2020, the mean daily cases in Malaysia was 55, while the highest daily cases of 277 was recorded on 6^th^ April 2020. During the period from 13^th^ March 2020 to 15 July 2020, the mean daily cases in Sarawak state in Malaysia was 5, while the highest daily cases of 32 was recorded on 1^st^ April 2020. Figure 1 shows the total number of cases stratified by the states of Malaysia.

**Fig 1.**
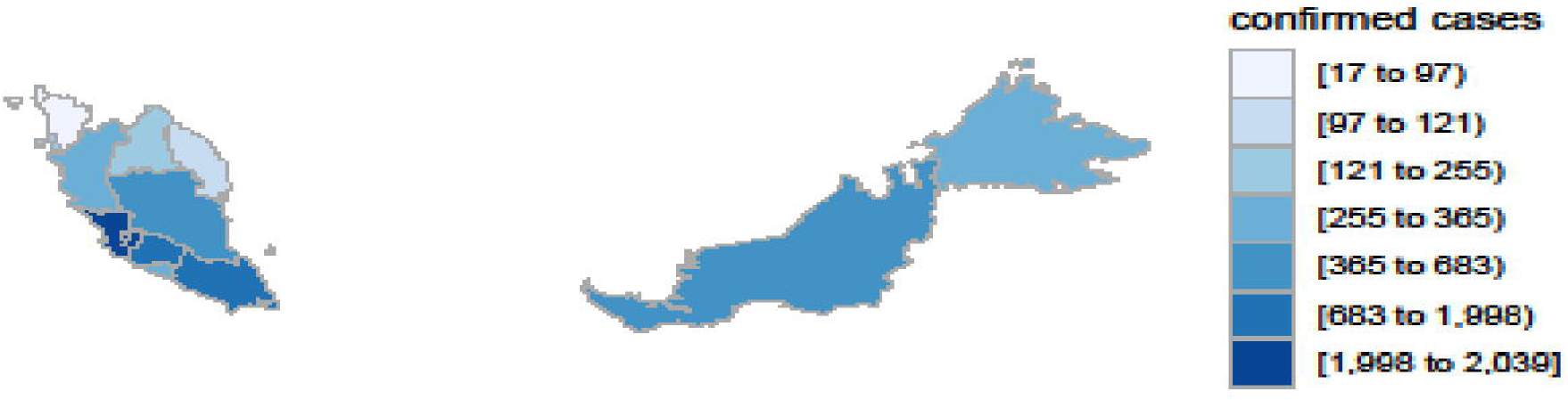
Total number of confirmed COVID-19 cases stratified by the states of Malaysia.

### Relationship with climate parameters

For Malaysia, the aggregate mean temperature, wind speed, precipitation and relative humidity were 89.8 °F, 5.3 mph, 6.1% and 99.4%, respectively (Table 1). The differenced daily COVID-19 cases in Malaysia over time from 24^th^ January 2020 to 15^th^ July 2020 are presented in Fig 2, which shows the time series data with an impression of stationary.

**Fig 2.**
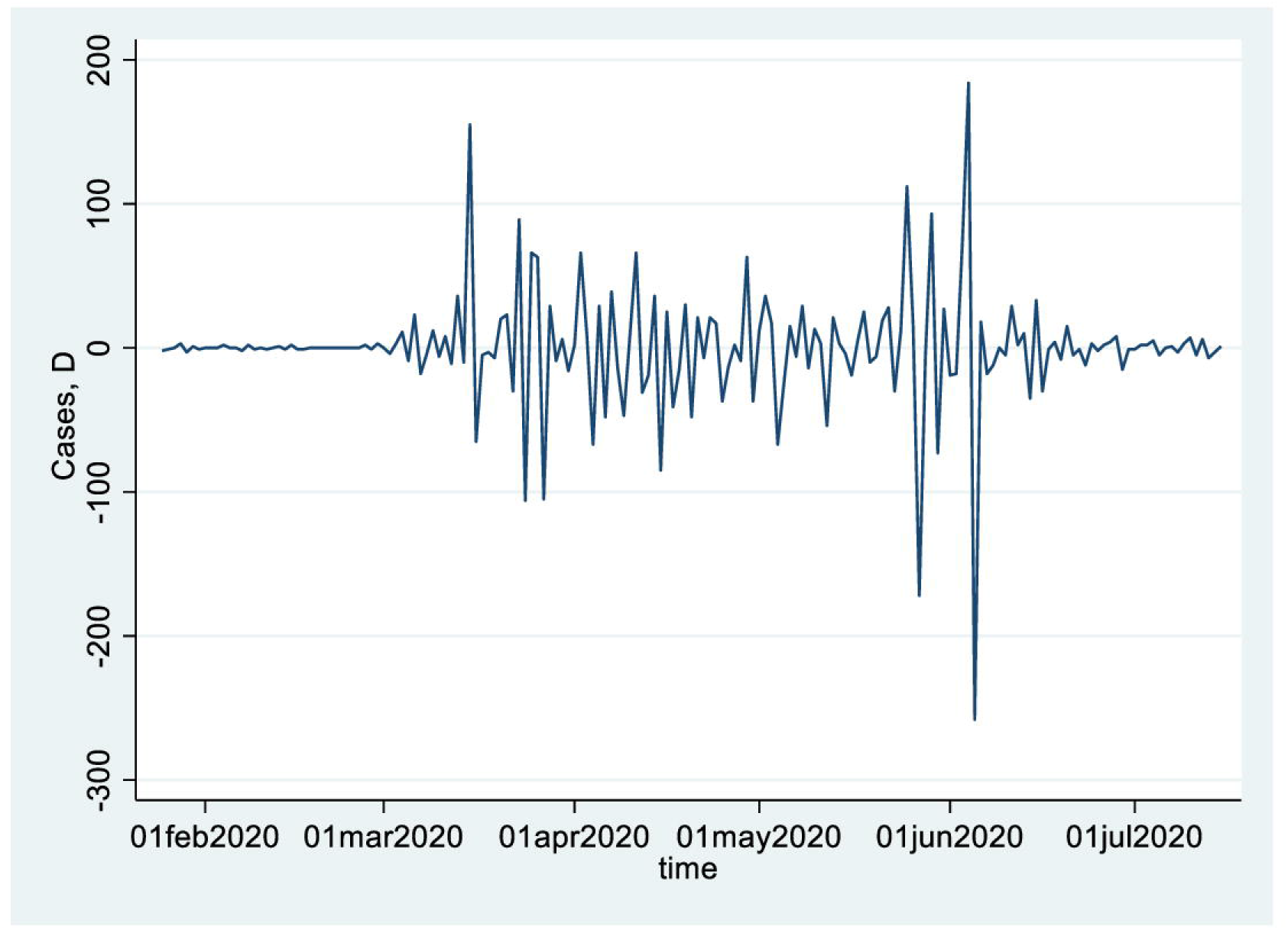
Differenced daily COVID-19 cases in Malaysia from 24 January 2020 to 15 July 2020.

The estimated coefficients of the ARIMA(1,1,1) model for Malaysia are presented in Table 2 Temperature (–58.99 (–97.6 to −20.4, *p* = 0.003 and wind speed (–77.03 95% CI:-221.5 to 67.5, *p* = 0.029). were significantly and inversely related to Covid-19 confirmed cases in Malaysia during the study period.

**Table 2.** Estimates of the regression coefficients of the ARIMA model for Malaysia between 24 January 2020 and 15 July 2020

| Parameter | Coefficient (95% CI) | *p*-value |
| --- | --- | --- |
| Intercept | -1.05 (-5.7 to 3.58) | 0.65 |
| Temperature (°F) | -58.99 (-97.6 to -20.4) | **0.003** |
| Precipitation (%) | 132.1 (-0.66 to 264.8) | 0.051 |
| Humidity (%) | 20.9 (-3.9 to 45.9) | 0.099 |
| Wind speed (mph) | -77.03 (-221.5 to 67.5) | **0.029** |

The partial autocorrelation function PACF) plot (S1 Fig) had a significant spike only at lag 1, reflecting that all the higher-order autocorrelations are effectively explained by the lag-1 autocorrelation.

For Sarawak state from 13^th^ March 2020 to 15^th^ July 2020, we followed the same procedures performed for the model of Malaysia (S2 Fig). None of the climate variables were significantly associated with the number of confirmed COVID-19 cases (Table 3).

**Table 3.** Estimates of the regression coefficients of the ARIMA model for Sarawak state of Malaysia

| Variable | Coefficient (95% CI) | *p*-value |
| --- | --- | --- |
| Temperature (°F) | -0.79 (-5.9 to 4.3) | 0.76 |
| Precipitation (%) | -0.16 (-19.0 to 18.7) | 0.99 |
| Wind speed (mph) | -7.02 (-26.8 to 12.8) | 0.49 |
| Constant | -0.04 (-0.37 to 0.3) | 0.83 |

### Forecasting of the confirmed COVID-19 cases

For Malaysia, ARIMA forecasting with the *auto.arima(*1, 1,1) function showed the best order of an ARIMA process with the appropriate degree of differencing and identification of the AR and MA parameters. This finding was also applicable to the *auto.arima*(1,1,1) model for Sarawak state. The forecast results for new daily COVID-19 cases in Malaysia and Sarawak state (prediction of 14 days) are presented in Fig 3 and Fig 4, respectively. We produced a final set of forecasts and prediction intervals using the most recent data, up until 15^th^ July 2020. We estimated two levels of uncertainty (80% and 95% CIs). Ljung–Box tests showed that the null hypothesis of the serial independence of the residuals for the current time series models could not be rejected (*p*> 0.63 for Malaysia, *p*> 0.87 for Sarawak state).

**Fig 3.**
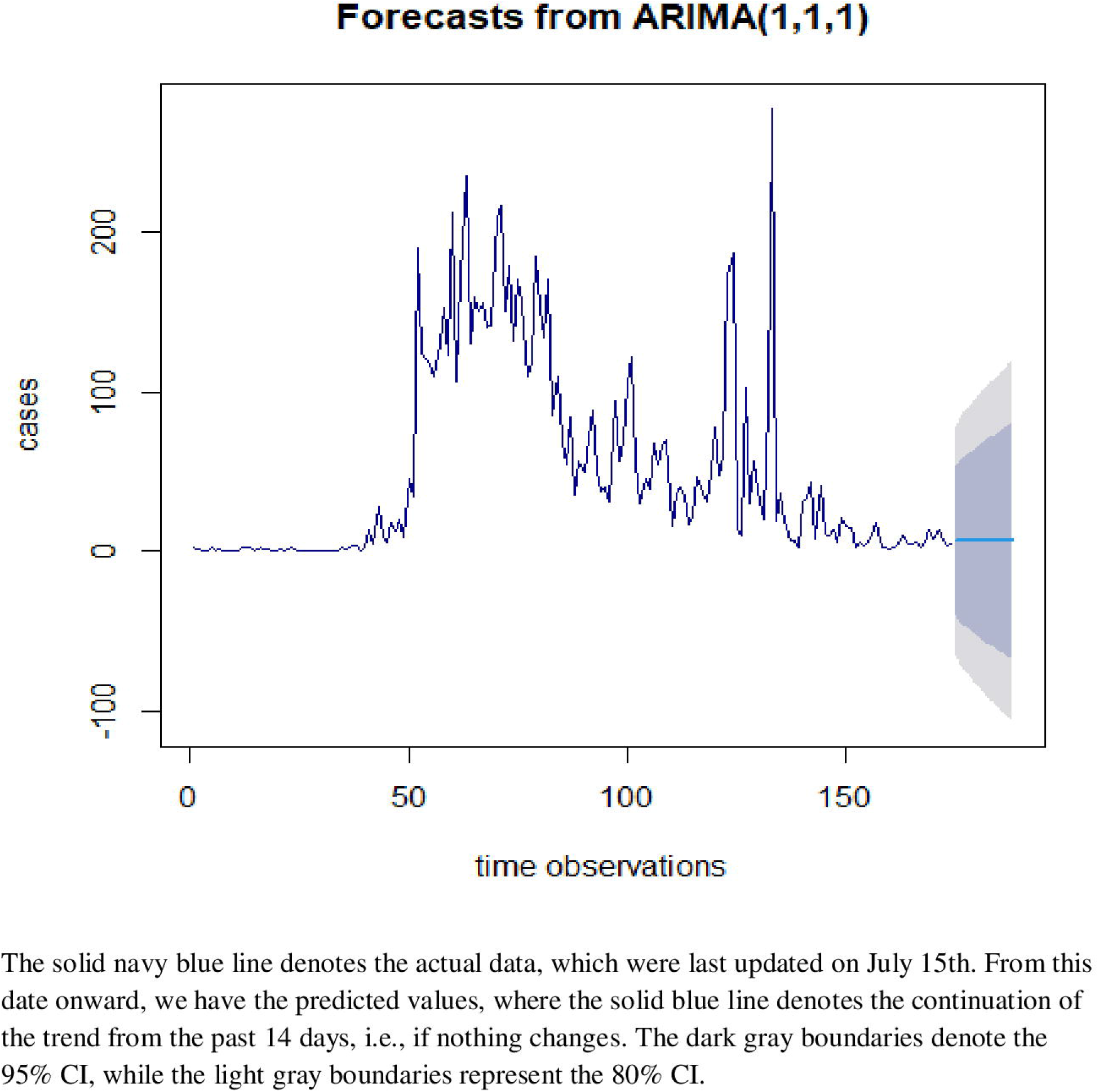
Forecasts of new daily COVID-19 cases in Malaysia (prediction of 14 days).

**Fig 4.**
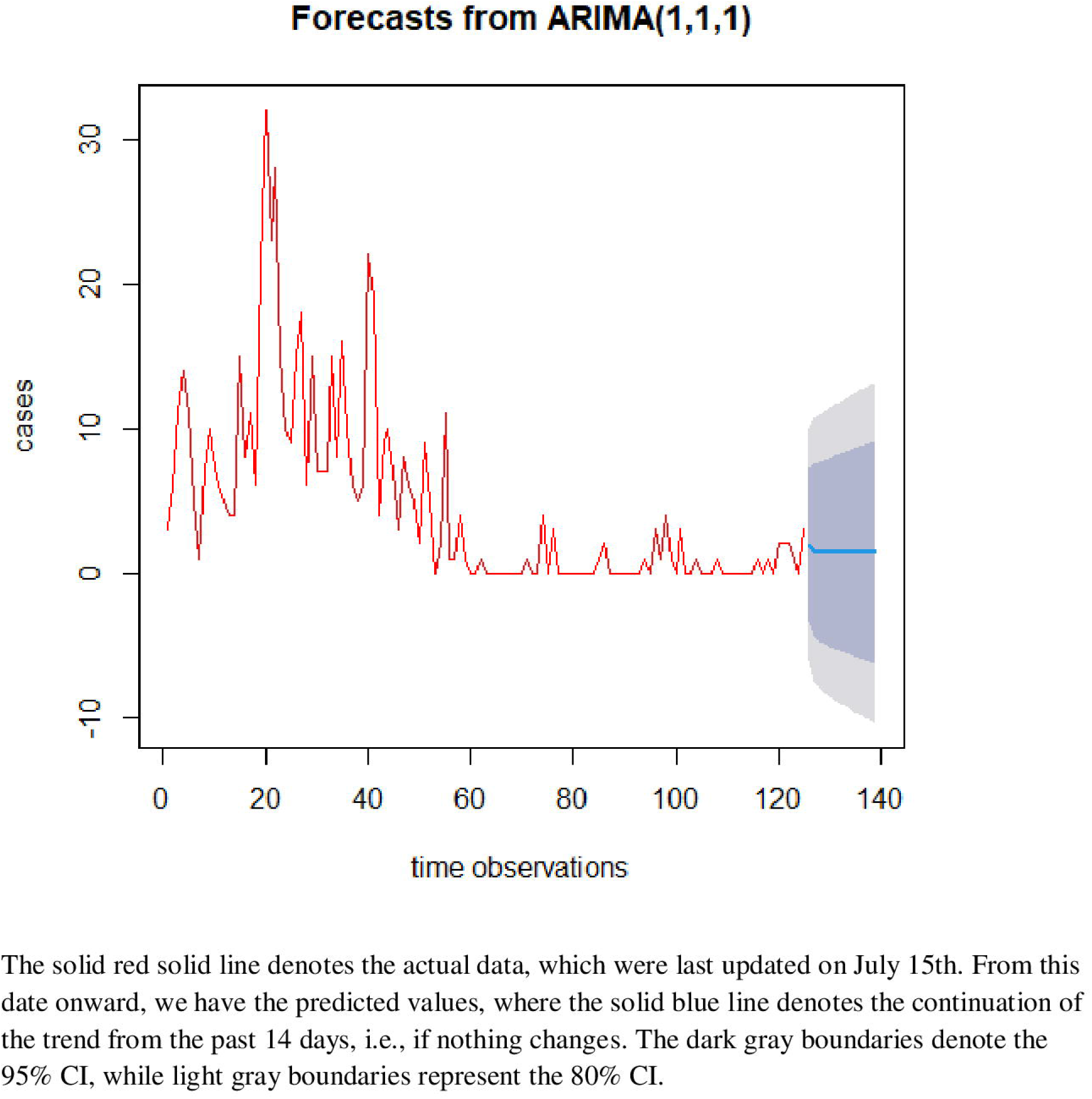
Forecasts of new daily COVID-19 cases in Sarawak state (prediction of 14 days).

As seen in the figures, the spreads were characterized by a declining trend, indicating that the COVID-19 transmission has probably already reached its peak. However, the slowdown in the new COVID-19 cases seems to require shorter time for Sarawak state compared to the whole country. On the other hand, a small spike (around a single digit) was seen in Sarawak state; this finding cannot rule out the possibility of a new wave (with small intensity) if the current recommendations/standard operating procedures such asocial/physical distancing, the use of face masks, and hand washing, as issued by the Malaysian MOH is not followed properly. The forecast accuracies were validated with the MAE, MAPE, MASE, and RMSE (S1 Table).

## Discussion

### Summary of the main results

The findings of this study were based on COVID-19-related data and climate parameters available for Malaysia in aggregate and Sarawak state in particular. The major observations of the current analysis are as follows:

1. For both Malaysia (24^th^ January 2020 to 15^th^ July 2020) and Sarawak state (13^th^ March to 15^th^ July 2020), time series ARIMA models were introduced.
2. For Malaysia, the findings suggest that, *ceteris paribus*, as the average temperature or wind speed increased, the number of COVID-19 cases decreased.
3. None of the climate parameters showed a significant relationship with the number of COVID-19 cases in Sarawak state.
4. Forecasts from the ARIMA models showed that new daily COVID-19 cases have already reached the outbreak level, and a decreasing trend was observed in both settings. C*eteris paribus*, a small number of new cases (approximately a single digit) would be indicative of a probable second wave in Sarawak state.

Our findings were supported by a study in Turkey, which also showed an inverse relationship between temperature and the number of COVID-19 cases [7].

A study in Singapore reported that an average maximum temperature increase of 1 °C decreased the incidence rate by −7.5% (95% CI = [−12.3; −2.6]) on the same day [8]. In the 2003. SARS epidemic, a study of Beijing and Hong Kong showed an inverse relationship between temperature and the number of cases [9]. Although an exact cause is not yet known, a possible reason may be that higher temperature influences the viability of the severe acute respiratory syndrome coronavirus 2 virus [7]. A higher wind speed was significantly and positively related to the number of new COVID-19 cases reported in a study in Turkey [7], which was contradictory to the current findings in Malaysia.

Temperature is a fundamental factor in the human living environment that can play an important role in public health in terms of epidemic development, prevention, and control [10]. Unpublished studies have also reported that low temperatures are beneficial to viral transmission [11,12].

Bill Gates commented on COVID-19 transmission saying that *“I hope it’s not that bad, but we should assume it will be until we know otherwise”* [13]. It is believable that forecasts and their associated uncertainty can and should be an integral part of the decision-making process, especially in high-risk cases [14]. In this study, univariate time series models which assume that the data are accurate and past patterns will continue to apply were used [15]. Significant, consistent forecast errors (potentially spanning outside the prediction intervals) should be associated with changes in the observed patterns and the need for additional actions and measures encountered.

The modeling quality is dependent on whether the assumptions are violated or not. In this study, the model’s diagnostic tests and a small percentage of error terms ensure the forecasting validity. The assumptions required for the prediction intervals of ARIMA models, such as uncorrelated and normally distributed residuals, were not violated. Moreover, the accuracy, robustness, coherency and uncertainty mitigation that the models imply have made them a popular forecasting framework, highlighting the potential benefits of multiple temporal aggregation in decision making [16]. Forecasts of deaths and hospitalizations (the number of new cases in this case) will help inform public health decision making by projecting the likely impact of the COVID-19 pandemic.

### Study limitations

There were limitations to the current analyses that need to be acknowledged. As the results are based on aggregate data for the country and specific to only one particular state, in light of geographic differences, the evaluation power could have been affected. Additionally, the results may have been affected by the quality of the data in relation to the accuracy of the diagnostic tests for COVID-19. Furthermore, there might be other factors (e.g., the quality of the case surveillance system) other than weather variables that can influence the transmission potential of COVID-19. Only a few weather variables were available for inclusion in the current study.

Nevertheless, there are strengths in the current study. The source data for COVID-19 cases originated from the national database, which is identical to the John Hopkins’s database, thus ensuring the accuracy of the data. The findings were, to a certain extent, comparable to those from other published studies. For instance, an inverse relationship between the average temperature and the number of new cases was in line with the findings from other countries. As we hope the presented forecasts clearly show, epidemic growth is a highly nonlinear process, where every day lost to inaction is too much [17]. On the other hand, a small spike (approximately a single digit) was seen in Sarawak state; this finding cannot rule out the possibility of new (small intensity) waves. This finding implies a potential small wave is expected in Sarawak state, if people fail to comply with the measures recommended by the Malaysian MOH such as social/physical distancing, use of face masks and hand washing. In this context, forecasts will be a useful tool for governments and individuals to make decisions and take appropriate actions to contain the spread of the virus [15].

## Conclusions

The findings suggest that it is important to consider climate parameters and forecasts to reduce the uncertainty in the severity of future COVID-19 transmission. Our results highlight that forecasts will be a useful tool for making decisions and taking the appropriate interventions to contain the spread of the virus in the community.

## Data Availability

The availability of all data in the manuscript and supporting information.

## Acknowledgments

We thank our institutions for allowing us to perform this study. We acknowledge Dr. Norah Htet for assisting with the earlier draft.

## Supporting information

S1 Fig Autocorrelation (ACF) and partial autocorrelation of COVID-19 time series data in Malaysia

S2 Fig Autocorrelation (ACF) and partial autocorrelation of COVID-19 time series data in Sarawak state

S1 Table Accuracy measures

